# Development of an early warning system for Nipah outbreak prevention: on-site inactivation, PCR surveillance and sequencing in Bangladesh

**DOI:** 10.64898/2026.03.17.26348576

**Authors:** Md Nurul Islam, Shahneaz Ali Khan, Zsófia Lanszki, Ágota Ábrahám, Sazeda Akter, Abdulla Al Mamun Bhuyan, Brigitta Zana, Md Shafeul Islam, Safia Zeghbib, Krisztina Leiner, A S M Rafsan Jani, Md Jalal Uddin Sarder, Md Hemayatul Islam, Nitish C Debnath, Johnny A Uelmen, Krisztián Bányai, Gábor Kemenesi, Sharmin Chowdhury

## Abstract

**Background:** Mobile laboratory diagnostic technologies for Nipah virus outbreak prevention, mitigation and response remain limited, despite the critical need for such capacities in remote, low-resource regions where most cases occur. We aim to address this gap by implementing a workflow that includes method development, laboratory validation, and field demonstration of a mobile Nipah virus complex diagnostic solution.

**Methods:** We developed a flexible mobile laboratory workflow incorporating PCR capacity, a novel amplicon-based sequencing protocol, and a validated Nipah virus inactivation procedure. Following development and validation, we demonstrated the feasibility of this workflow through repeated field sampling of bat colonies in Nipah virus endemic regions of Bangladesh across multiple field campaigns.

**Findings:** We demonstrated the feasibility of this system for early outbreak response and as a potential early warning tool prior to the emergence of human cases. We detected two urine samples from flying foxes that tested positive and performed full-scale on-site analysis, including qPCR diagnostics and NGS sequencing, within 24 hours.

**Interpretation:** As highlighted in the present study, active surveillance enables outbreak prevention by identifying bat colonies that are actively shedding viruses in real time, even in rural settings. Also, this method can provide rapid, on-site sequence data to track and better understand the genomic diversity of Nipah virus in natural reservoirs during both outbreak and non-outbreak periods. In this study we aimed to establish the foundations of a standard procedure for safe and rapid field testing of Nipah virus in remote areas.

**Funding:** This study was funded by the National Research, Development and Innovation Fund of Hungary – Grant details: Strengthening of Regional Research Infrastructure, Capacity Building and Pandemic Preparedness (2024-1.2.3-HU-RIZONT-2024-00041).

**Research in context:** *Evidence before this study:* We searched the PubMed database with the following search term without fixed timeframe: (“Nipah virus” OR Nipah) AND (Bangladesh OR India OR Southeast-Asia) AND (“mobile laboratory” OR “field laboratory” OR “point-of-care” OR portable OR onsite) The search yielded 12 publications that address specific elements of mobile laboratory solutions and provide opinions on the necessity of mobile technologies (point-of-care testing, rapid diagnostics). One technological paper was identified in relation to a bus-based mobile laboratory solution in India, which is also referenced in our manuscript. Further manuscripts are related to the molecular and clinical diagnostics of Nipah virus, and a genome sequencing protocol validated on clinical samples was also identified. The health risk posed by Nipah virus can be reduced by investments in research, development and technology, as described in the literature. However, the available techniques can currently be interpreted as separate modules and a comprehensive diagnostic method validated in terms of safety, complex technical solutions and usability in real world conditions is still lacking.

*Added value of this study:* This complex study, which ranges from laboratory development to validation of methods and demonstration in real-world conditions, is the first of its kind. There is currently a limited number of methodologies in the literature and a significant need, which is also included in the WHO regional Nipah virus epidemic prevention plan. The development of rapid on-site surveillance techniques, starting from validated virus inactivation protocol, through the on-site real-time RT-PCR screening and amplicon-based sequencing method represents a significant advancement in early detection and monitoring of NiV. This approach facilitates swift responses in the early stages of human cases and contact tracing strategies or identification present of virus in bat colonies even before human cases emerge, bolstering outbreak prevention efforts.

*Implication of all the available evidence:* Our study highlights the need for a critically important technology integration. As the use of mobile laboratories expands and the demand for rapidly deployable, mobilizable technologies grows, there is an increasing need to move toward standardized operational approaches. The first step is to test different technologies in real-world situations beyond the development phase and provide alternatives for use during or in the prevention of epidemic situations.

## Introduction

Nipah virus (NiV) is a bat-borne zoonotic virus that can cause respiratory distress and fatal encephalitis with high case fatality rates in humans^1^. It is recognized as a WHO priority pathogen for research and development. Several outbreaks among humans and livestock have occurred since its discovery in Malaysia in 1999 ^2,3^. Following the Malaysian outbreak, flying foxes (*Pteropus* spp.) were hypothesized as the natural reservoir for Nipah virus. Years of surveillance efforts among Indian flying foxes in Bangladesh supported this hypothesis, and *Pteropus medius* are now considered the natural reservoir for NiV. The distribution range of these bats across major parts of South and Southeast Asia suggests a greater risk of human infections with NiV. This risk has been validated by the emergence of human cases in India over the last few years ^4,5^. Outbreaks among humans have been reported in multiple countries so far, but most cases are still reported from Bangladesh, and across countries, most are affecting densely populated rural areas. Most of these cases and outbreaks are epidemiologically linked to the consumption of raw date palm sap during the winter season, which can be contaminated by visiting bats during the collection process on the trees. Although additional transmission routes are hypothesized, investigating them warrants further research ^6^.

The emergence of the virus in humans in multiple regions within the distribution area of the natural reservoir flying foxes indicates the growing importance for outbreak preparedness and the development of effective early warning systems. Although viral spillover is rare, it most often occurs in remote areas with limited diagnostic capacity. The availability of rapid diagnosis is a key element in effective outbreak control. Based on currently available literature, the outcome of Nipah infection varies greatly depending on the strain causing the infection (i.e., asymptomatic infections are known with the Malaysia strain, whereas they are rare or absent with the Bangladesh strain). Rapid diagnosis can be useful in all scenarios, whether it is for epidemic management or for exploring epidemiological background factors ^7^. Based on available epidemiological data, Bangladesh is currently the global hotspot for most human cases. Since 2001, a total of 341 cases and 241 deaths were reported (71% fatality rate), mostly in association with annual sporadic cases and some localized outbreaks ^8,9^.

Developing novel diagnostic platforms - including point-of-care molecular tools and field-deployable assays - can significantly improve case identification in remote areas where spillover most often occurs. Integrating these tools into surveillance networks, supported by community engagement and real-time data sharing, will accelerate outbreak recognition and intervention, a priority aim in the recent Nipah outbreak prevention and control strategy ^10,11^. Such technological integration and mobilization can ultimately shift the fight against Nipah from epidemic investigation to early detection and the establishment of an early warning system.

Even with efforts to understand and mitigate Nipah virus transmission, challenges remain, as demonstrated by the recent outbreak in India in 2026 ^10,12^. The precise origin remains unclear, underscoring the need for continued research and surveillance to better understand Nipah virus transmission dynamics and improve outbreak preparedness and response measures in the region. The current Nipah epidemic also highlights the important role of mobile laboratory capabilities, as the Indian public health authority deployed one in this most recent outbreak ^10,13^.

The need for early detection or early warning systems and rapidly deployable laboratory solutions has already been noted in several professional publications. The complex field-adapted workflow we present in this study can be suitable for environmental or bat monitoring before epidemics, for data generation and analysis during epidemics, and for rapid epidemic management steps. The advantage of this approach is the potential to serve neglected or rural regions without adequate diagnostic capacity and to foster rapid outbreak identification and control in these areas. To our knowledge, this study represents the first end-to-end, field-validated protocol for NiV - spanning on-site virus inactivation, qPCR, and nanopore sequencing - deployed directly in a low-resource endemic setting.

## Methods

### Inactivation of infectious Nipah virus

Cytotoxicity testing was performed using the VERO-E6 (ATCC, USA) cell line prior to the inactivation experiment to determine the concentrations of commercial lysis buffers that could be tolerated by the cells. Based on these results, subsequent experiments allowed the dilution of the inactivated sample–lysis buffer mixtures to concentrations that were non-cytotoxic to the cells.

For the cytotoxicity testing, 35000 VERO-E6 cells per well were seeded in 96-well plates in DMEM (Dulbecco’s Modified Eagle Medium) supplemented with 10% FBS (Fetal Bovine Serum) and 1% PS (Penicillin-Streptomycin).

For the experiment, serial dilutions were prepared from mock-inactivated samples (DMEM + lysis buffer) diluted in DMEM (2% FBS, 1% PS). Three rounds of cytotoxicity tests were conducted. In the 1^st^ round, dilutions with bigger steps were used to determine the approximate dilution range that is tolerable for the cells. In the 2^nd^ round smaller steps of dilutions were used in the range defined by the 1^st^ round results. The last round was to precisely define the optimal dilution that is not too diluted but still tolerated by the cells. Figure 1. summarizes the main steps of the cytotoxicity testing workflow.

**Fig. 1:**
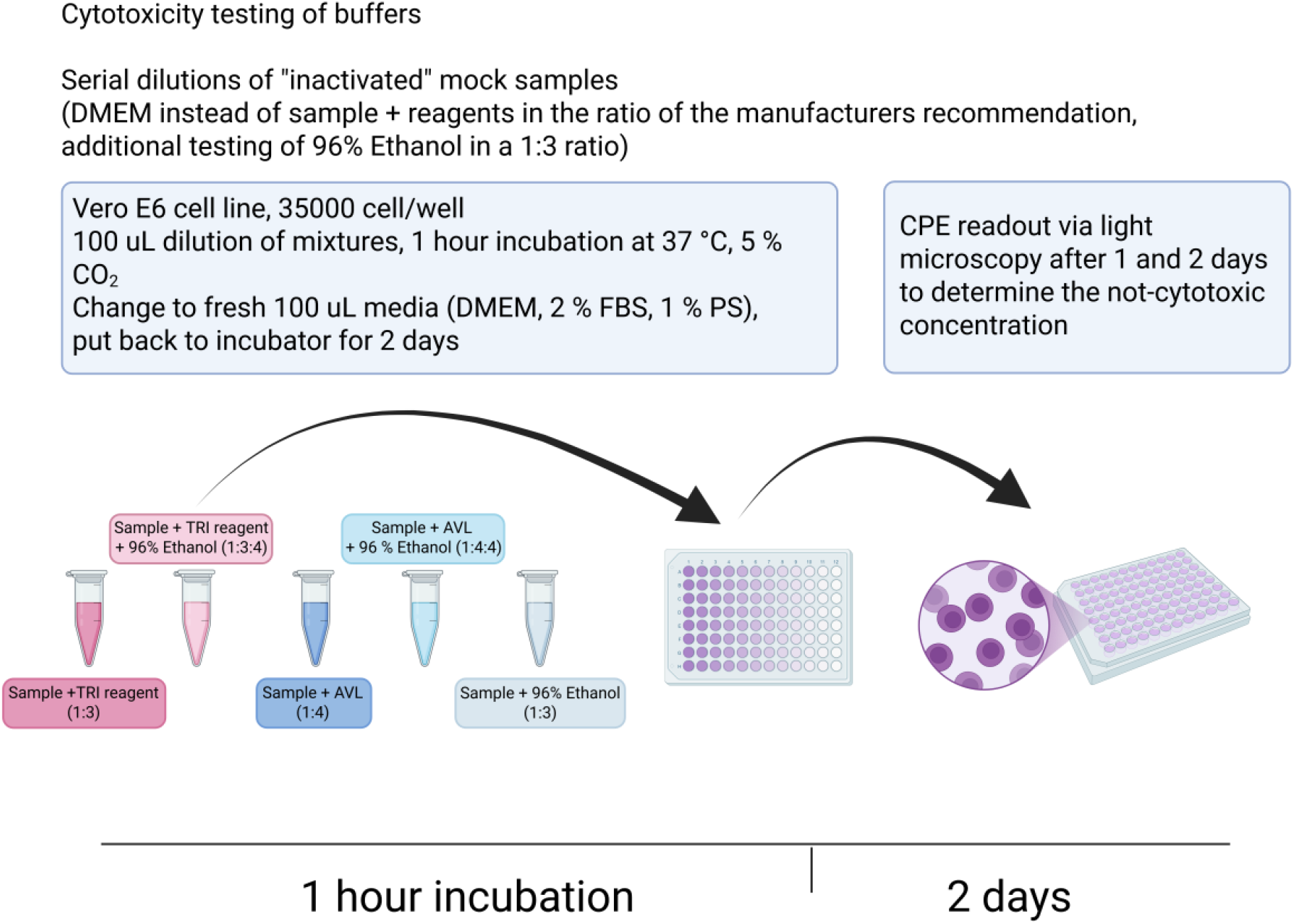
Flow chart of the buffer cytotoxicity testing *in vitro* prior to inactivation experiments with live Nipah virus

All work with infectious Nipah virus isolate were carried out in the BSL4 laboratory of the National Laboratory of Virology, University of Pécs, Hungary. Inactivation of Nipah virus (Malaysia strain) was carried out with a stock with 1×10^5^ TCID50 virus titer. Inactivation was performed according to the manufacturer’s instructions, using buffer concentrations and incubation times as specified, to fully comply with normal use conditions. Also, inactivation with only ethanol was performed the same way as with TRI-Reagent (1:3 sample-to-buffer ratio and no incubation time). VERO-E6 cells were seeded on 6-well plates with 750000 cells/ well in DMEM supplemented with 10% FBS and 1% PS. Samples inactivated with any buffer were diluted to a final volume of 10mL (as previously determined during the cytotoxicity experiments) and pipetted into 5 wells (2 mL each) for the duration of the treatment. With this approach, the whole inactivated liquid volume was used for the treatment of the cells to fully validate the inactivation.

On each 6-well plate, 1 well was used as a negative control and 5 wells for the samples. For the positive control 3 different dilutions were used, one corresponding to the least diluted inactivated sample, one to the most diluted inactivated sample and one in the mid-point on a separate plate. A negative control well was also used on the positive control plate. DMEM, 2% FBS, 1% PS was used as the negative control during inoculation. After 1 hour incubation with the inoculation liquids, we changed to fresh media (DMEM, 2% FBS, 1% PS), 4mL/well. After 4 days, plates were scanned with a CytoSmart device (Axion Biosystems, Netherlands) and assessed for CPE, then kept frozen at −80 before proceeding to the next validation step.

For the next part of the experiment, samples were transferred to 24-well plates seeded with 350000 cells/well, with the same culture conditions as before. 400 µL from each sample of the first round of the experiment, was transferred to the 24 well plate to 6 wells (2400 µL in total). Negative and positive controls were similarly transferred to the new plate. These samples were incubated for an additional 4 days at 37°C in 5% CO_2._ After 4 days, plates were scanned as previously and assessed for CPE, then frozen at −80°C. A summary of the inactivation experiment workflow is available (Fig. 1).

In addition to CPE monitoring, reverse transcription quantitative PCR of the supernatants was done to compare viral genomic RNA amount at multiple sampling points from the inoculation. This allowed the monitoring of potential viral genomic RNA titer growth and added another validation layer to CPE monitoring. For PCR reactions, samples were taken from the starting point of the whole inactivation experiment, then after thawing the 6-well plates, and then after thawing the 24-well plates. From each sample (inactivated, positive control, negative control) 100 µL was taken and pipetted into a 2 mL screw cap tube, then 300 µL TRI-Reagent was used on each for sample inactivation. In the next step nucleic acid was isolated from the samples with Direct-zol RNA Miniprep kit (Zymo Research, USA). For the qRT-PCR the reaction was performed using the Brilliant III Ultra-Fast QRT-PCR kit (Agilent, USA). Nipah N gene specific primers and probe were obtained from IDT, and their sequence were previously published on Protocols.io^14^. Nipah specific primers were NiV-N-TM2018_For CTGGTCTCTGCAGTTATCACCATCGA, NiV-N-TM2018_Rev ACGTAYTTAGCCCATCTTCTAGTTTCA, and the probe was NiV-N-TM2018_Prb FAM-CAGCTCCMGACACTGCCGAGGA-BHQ1^14^. The amplification protocol consisted of reverse transcription at 50 °C for 10 minutes, followed by an initial denaturation at 95 °C for 3 minutes, and 40 cycles of denaturation at 95 °C for 5 seconds and annealing/elongation at 60 °C for 30 seconds, and the reactions were run on a MIC qPCR platform (Bio Molecular Systems, Australia).

### Field sampling of bat roost samples

To minimize bat disturbance during sampling, large polyvinyl sheets were placed under the bat roosts during the night, when *Pteropus* bats are active and before they return to their roosts (before 4:00-5:00 a.m.). After the entire colony returned to the roost, urine samples were collected directly from the foil using a 1 ml pipette and pooled in a 2ml collection tube containing TRI-Reagent. The TRI reagent proved to be a suitable inactivating agent in our preliminary experiments; therefore, the samples were collected directly into this buffer without exceeding the experimentally proven inactivation ratio of mixed sample/buffer. During pooling, the intention was not to mix more than 2-3 samples together. Plastic foils were cut in variable sizes between 1m x 1-3 meters pieces, depending on the characteristics of the landscape below the bat roost and with the intention to cover as much area as we can with separate and non-overlapping foil pieces. For personal protective equipment (PPE) we used full body covering tyvek suits, including neck and feet protection parts. As respiratory and face protection we used P3 filtered masks and face shields combined with protective glasses.

### Nucleic acid extraction and PCR

Samples were processed directly in the field after collection using an on-site mobile laboratory. RNA was extracted using the Direct-zol RNA MiniPrep (Zymo Research, USA) combined with RNase inhibitor treatment at elution (TargetEx RibonEx RNase inhibitor, TargetEx, Hungary). Subsequently, RT-qPCR screening was performed using the same PCR protocol as described for the inactivation of infectious Nipah virus. Thereafter, the MyGo Mini PCR system platform (IT-IS Life Science, Ireland) was utilized to complete the run, while MyGo PCR software (v.3.5.2) was used for sample analyses. The remaining nucleic acid samples were stored frozen.

### Sequencing method and bioinformatic analysis

We used PrimalScheme to generate a primer pool for sequencing, aligned to NiV sequence dataset from different stains and hosts, covering a set of 45 sequences (1 from Thailand, 1 from India and 43 from Bangladesh). Manual corrections and primer modifications were done in Geneious Prime® 2025.2.2 software. For initial validation we used the nucleic acid extract of the Nipah Malaysia strain from an isolate with a 1×10^5^ TCID50 infectious virus titer. The list of reference sequences for primer design and primer sequences with their genomic positions are included as a supplement (Supplementary Table S1, S2).

The entire genome sequencing was conducted using MinION nanopore sequencing technology from Oxford Nanopore Technologies (Oxford Nanopore Technologies, UK). An amplicon-based sequencing method was developed, inspired by established protocols ^15^. Briefly, for cDNA preparation from the NiV positive RNA sample, Superscript IV (Invitrogen, USA) was utilized alongside random hexamers. Genome-specific, overlapping amplicons were then generated from the cDNA using the Q5 Hot Start HF Polymerase (New England Biolabs, USA) employing multiple primers. The primer sequences used were designed to generate overlapping amplicons of approximately 600 and 900 nt in size. PCR products were purified using the AMPure XP beads (Beckman Coulter, USA). The end-repair and dA tailing processes were carried out using the NEBNext Ultra II End Repair/dA-Tailing Module (New England Biolabs, USA). Thereafter, barcodes derived from SQK-NBD114.96 (Oxford Nanopore Technologies, UK) were ligated using the Blunt/TA Ligase Master Mix (New England Biolabs, USA). Following this, the pooled barcoded samples underwent joint cleanup with Ampure XP beads, after which the NA sequencing adapters were ligated using the NEBNext Quick Ligation Module. The final library was quantified using the Qubit dsDNA HS Assay Kit (Invitrogen, USA) on a Qubit 4 fluorometer. The final library was loaded onto a R10.4.1 flow cell (Oxford Nanopore Technologies, UK) on MinION Mk1B.

After sequencing, the raw data was quality checked by Nanoplot (v1.32.1). Basecalling of raw data was performed by Guppy (ONT Guppy v6.5.7) using the super accuracy base-calling algorithm (dna_r10.4.1_e8.2_400bps_sup config file). Demultiplexing and trimming of barcodes were performed with Guppy by using the default parameters of the ‘guppy_barcoder’ runcode. Mapping the trimmed reads to reference was done using Minimap2 (v2.24)^16^ plugin in Geneious Prime® 2025.2.2 software (https://www.geneious.com). The generated consensus sequences from medaka analyses and from mapping by the Minimap2 were aligned using the MAFFT plugin ^17^, and the base-calling errors were manually corrected in Geneious Prime® 2025.2.2 software.

### Phylogenetic analysis

The maximum likelihood phylogenetic tree was constructed in the IQ-TREE software package v3.0.1. ^18^. For phylogenetic analysis, we used the entire partial nucleotide sequence available from amplicon sequencing and all complete NiV genomes available in GenBank depository. GenBank database was accessed on 20.01.2026. Nucleotide sequences were aligned using MAFFT plugin ^17^ built in Geneious Prime® 2025.2.2 software. For the selection of the best-fit model, we used the ModelFinder implemented ultra-fast model selection feature of IQ-TREE software package. The tree was generated according to the suggested best-fit model GTR+F+R2. During analysis, we used the Bootstrap resampling method with 1000 replicates.

The maximum likelihood phylogenetic tree of the short nucleoprotein sequences of NiV was built by the same methodology as described previously, but the applied best-fit model was K3P+G4. The constructed trees were visualized and edited in the iTOL online tool ^19^.

### Haplotype analysis

For haplotype analysis of our short NiV sequence (136 bp) from 2022 all NiV nucleoprotein sequences were collected from GenBank database accessed on 20.01.2026. MUSCLE alignment and trimming of sequences were done by using Geneious Prime® 2026.0.2 software package. The haplotype file was generated in the DnaSP v6 program. Haplotype analysis was carried out in PopART 1.7 software.

### Ethical considerations

All fieldwork activities and non-invasive roost sampling of flying fox colonies were approved by the Bangladesh Forest Department, Ministry of Environment and Forest (Memo No: 22.01.0000.000.101.23.0018.23.1246). Because sample collection was restricted to environmental sampling of urine from underlying surfaces—designed specifically to prevent animal disturbance and eliminate the need for capture or handling—formal approval from an institutional animal care and use ethics committee was not required.

## Results

### Validation of inactivation procedures for live Nipah virus

Inactivation of live Nipah virus by commercially available nucleic acid extraction buffers was validated under BSL-4 laboratory conditions at the University of Pécs, Hungary. Virus inactivation was carried out according to the manufacturer’s protocol, using the supplied lysis buffers. All samples were confirmed to be fully inactivated by observation over 4 days post-infection with multiple passages, during which no cytopathic effect was detected. In addition to visual observation, PCR analysis provided supporting data from three sampling points: 0 DPI (days post-infection) and 4 DPI (stop-point). After a further passage, a third sampling was done at 4 DPI. In contrast to this, all positive controls produced strong CPE on the cell monolayers by day 4.

Based on the validation experiment results, TRI Reagent inactivated the Nipah virus sample upon addition. This is consistent with the manufacturer’s recommended protocol, which does not require an incubation time. Interestingly, TRI Reagent alone showed inactivation ability before the addition of EtOH, making it particularly suitable for rapid inactivation during sample collection in the field. For clarity, the experimental flow and main results are summarized on (Fig. 2).

**Fig. 2:**
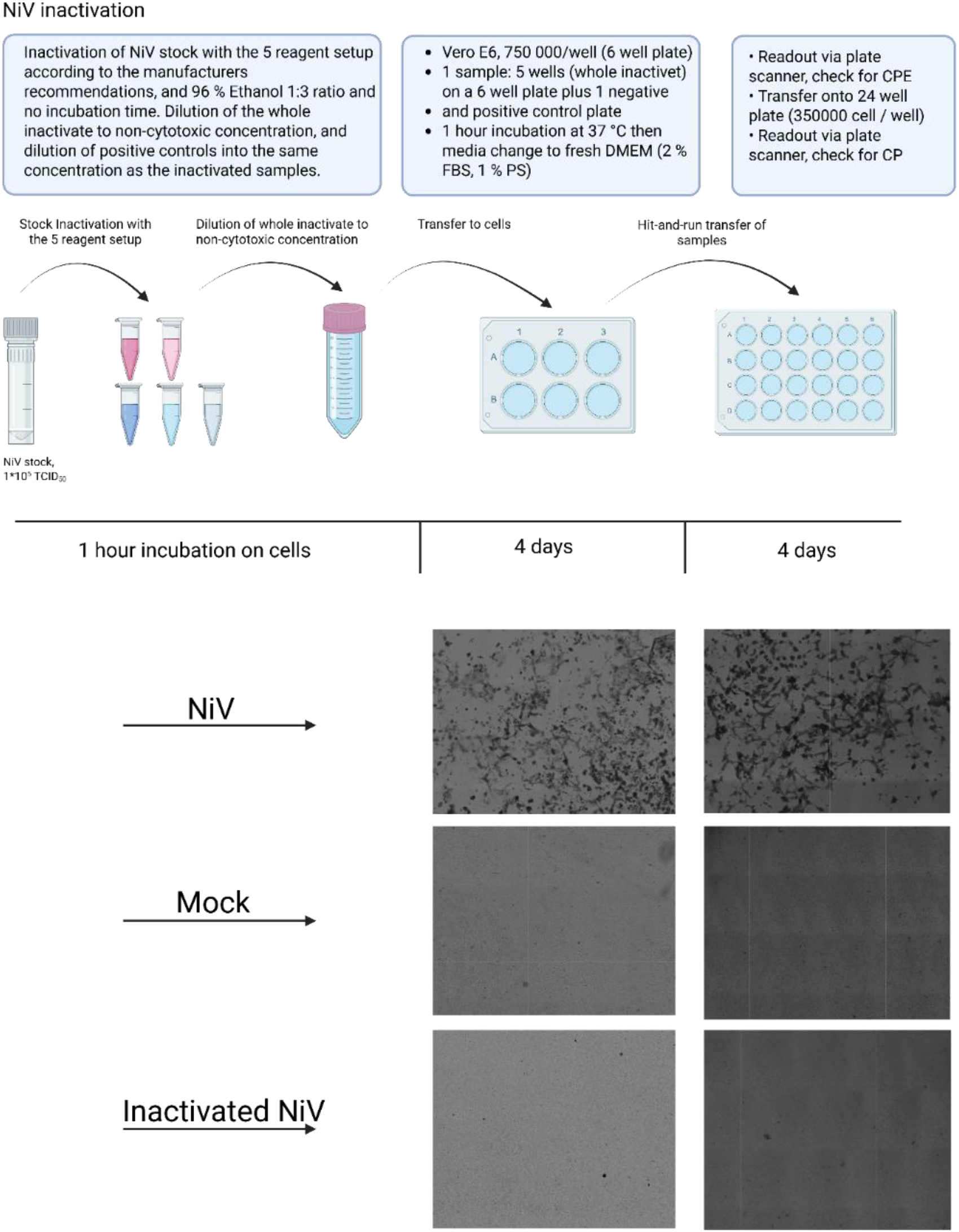
Flow chart of the live Nipah virus inactivation experiment *in vitro* with experimental details step-by-step.

### Sequencing method development

We designed an amplicon sequencing primer pool with dual strategy to amplify two set of amplicons with different sizes (600 and 900 bases pairs). The *in silico* development intended to design a system which is specific to a diverse set of Nipah Bangladesh strains. We were able to validate the test by sequencing the Nipah virus Malaysia strain and a field-collected roost urine sample from bats from Bangladesh. As a result of the sequencing run, we were able to reach an average coverage of 24760x, 24113x, 1697x and 239x per base with the Malaysia Nipah strain serial dilutions and a 3971x per base with the sample from Bangladesh. Genome completeness reached 97, 89, 61, 54 % in case of the dilutions from the Malaysia strain isolate and 30 % with the raw sample from Bangladesh (Figs. 3 and 4). More detailed quality attributes for each sequencing run are available in the supplementary materials for each sequencing run (Supplementary Table S3). Copy number calculations are available for all tested samples, along with the calculation background in the supplementary materials (Supplementary Figure S1, Supplementary Table S4-S6).

**Fig. 3:**
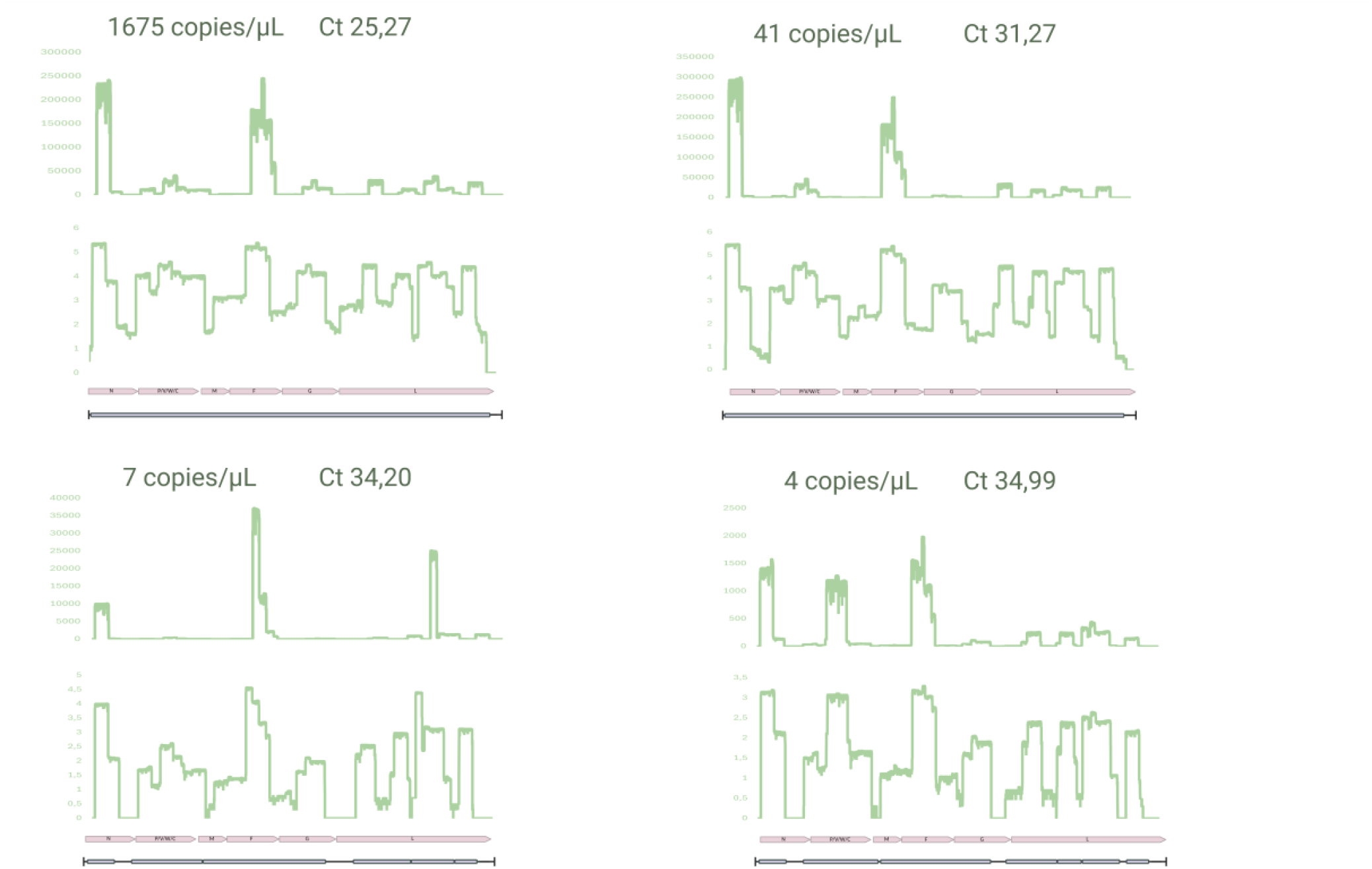
Sequencing coverage plots for the sequencing validation experiments, using the Nipah virus Malaysia strain in different dilutions. Visualization of sequencing coverage obtained using the NiV-specific amplicon-based sequencing method. Horizontal axis represents the genomic position, whilst the vertical axis displays the coverage values of the sequencing reaction.

**Fig. 4:**
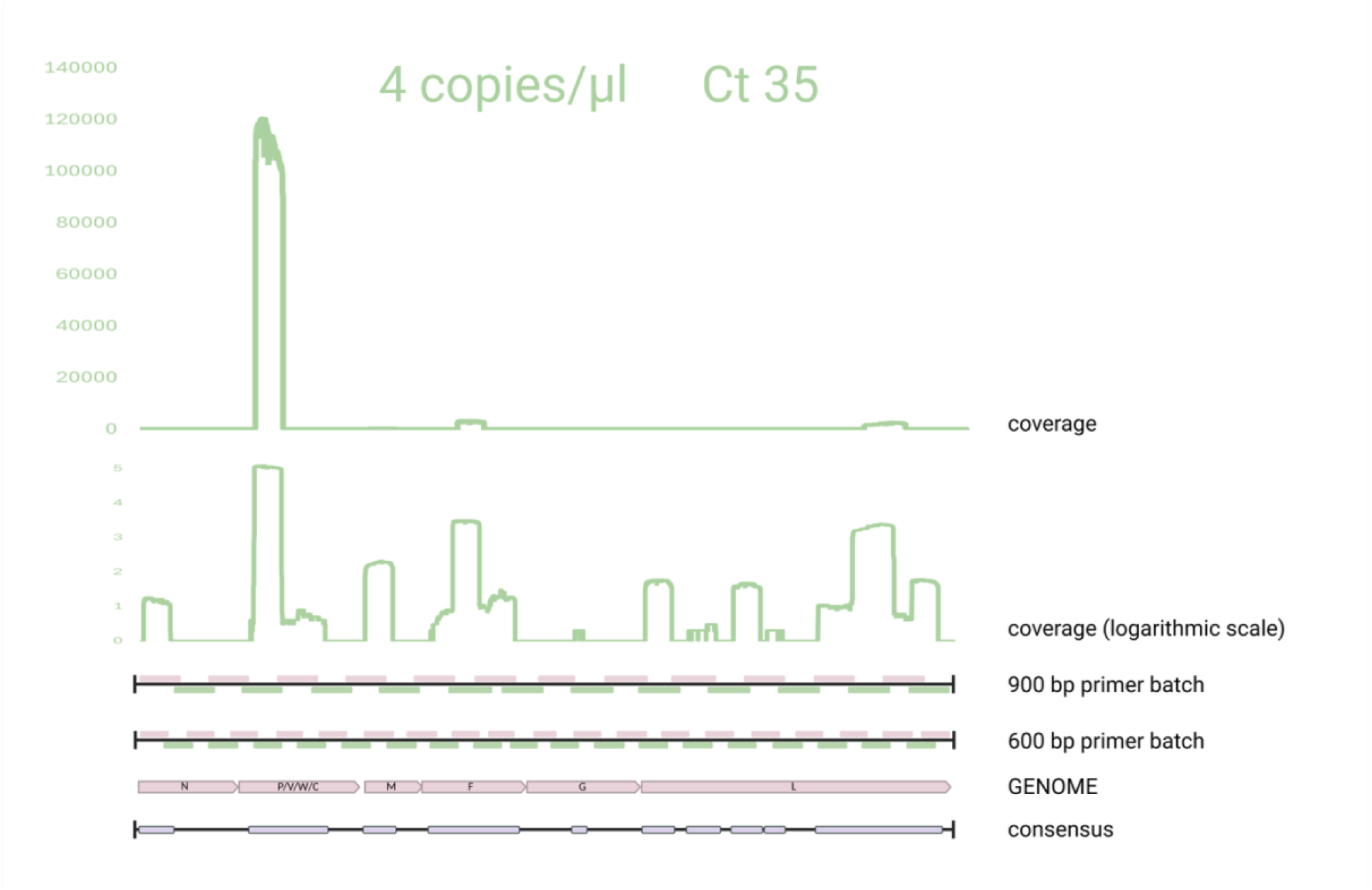
Sequencing coverage plot of the field collected positive bat roost urine sample. Visualization of sequencing coverage obtained using the NiV-specific amplicon-based sequencing method. Horizontal axis represents the genomic position, whilst the vertical axis displays the coverage values of the sequencing reaction.

### Sampling and NiV surveillance

To test the developed workflow under realistic conditions, we collected flying fox colony samples using non-invasive methods during multiple different climatic seasons and over three years (Table 1). Field sampling was conducted during three distinct period. Samples were collected between 1 and 19 January 2022, between 16 and 23 February 2023, and between 9 and 19 August 2025, covering multiple seasons and sampling locations, but all within the Nipah belt region (Fig. 5).

**Table 1:** Summary of collected and tested samples per year and type of the sample.

|  | 2022<br>January | 2023<br>February | 2025<br>August | Σ |
| --- | --- | --- | --- | --- |
| pooled urine | 560 | 256 | 101 | 917 |
| guano | 39 | 1 | 14 | 54 |
| food droplet | 7 | 0 | 0 | 7 |

**Fig. 5:**
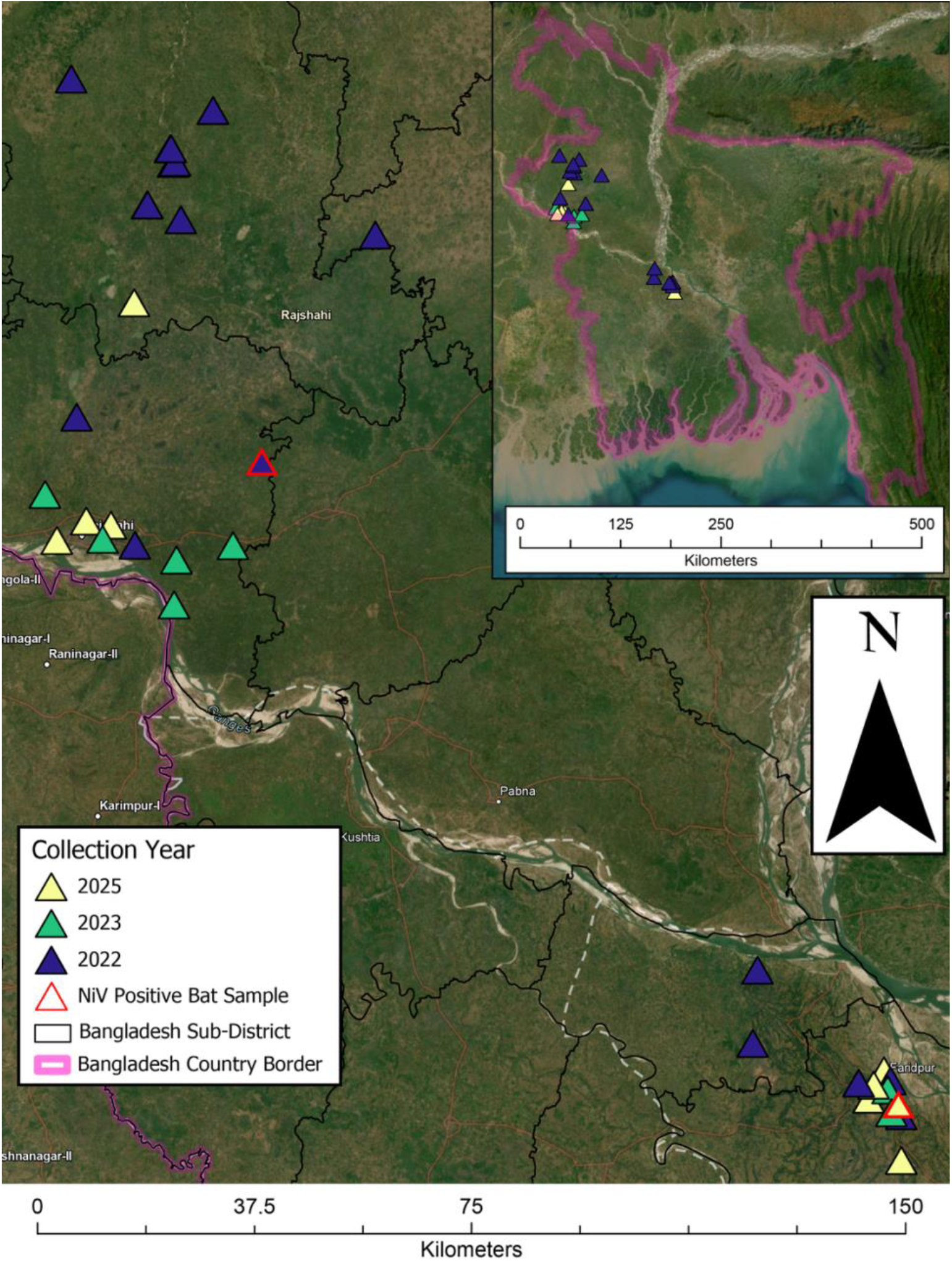
Spatial distribution of bat sampling sites in Bangladesh between 2022 and 2025. Triangles indicate sampling locations, colored by year of collection (blue: 2022; green: 2023; yellow: 2025). The red-outlined triangle marks the location of the Nipah virus (NiV) positive bat sample. Black lines represent sub-district boundaries within Bangladesh, while the pink line indicates the national border. The inset map shows the location of the study area within Bangladesh.

Of the 917 pooled urine samples tested, two were PCR positive, from 2022 and 2025 - Katakhali, Rajshahi district on January 17, 2022 and Faridpur district in Bangladesh on August 17, 2025. Based on the Ct values, the 2022 sample contained extremely low levels of NiV genomic RNA (ct greater than 40), but we were still able to successfully sequence the short amplicon of the real-time qPCR on-site, confirming the presence of the virus and demonstrating the possibility of field, laboratory-independent NGS sequencing. The Ct value of the urine sample from 2025 was 35, and on-site genome sequencing only amplified a few genomic regions, but from these, we were able to successfully analyse the genetic relationship of the virus with other strains. None of the guano or food droplet samples yielded a positive result.

### Phylogenetic analysis

The phylogenetic analysis shows that the strain detected in 2025 is grouped with the endemic Bangladeshi strains. Homology analysis of the 2022 sample using a shorter genome segment also yields a similar conclusion. Using the mobile diagnostic and genomic method we applied, we detected the endemic NiV strains directly from bat colony samples.

In the haplotype network, the sequence from 2022 is connected by a limited number of mutational steps, suggesting it is not highly divergent but is clearly distinguishable from the most prevalent haplotypes circulating in Bangladesh. The position of this sequence on the phylogenetic tree places it among sublineage 2 sequences (Fig. 6). The data used for haplotype analysis and their distribution by haplotype are part of the supplementary material (Supplementary Table S7).

**Fig. 6:**
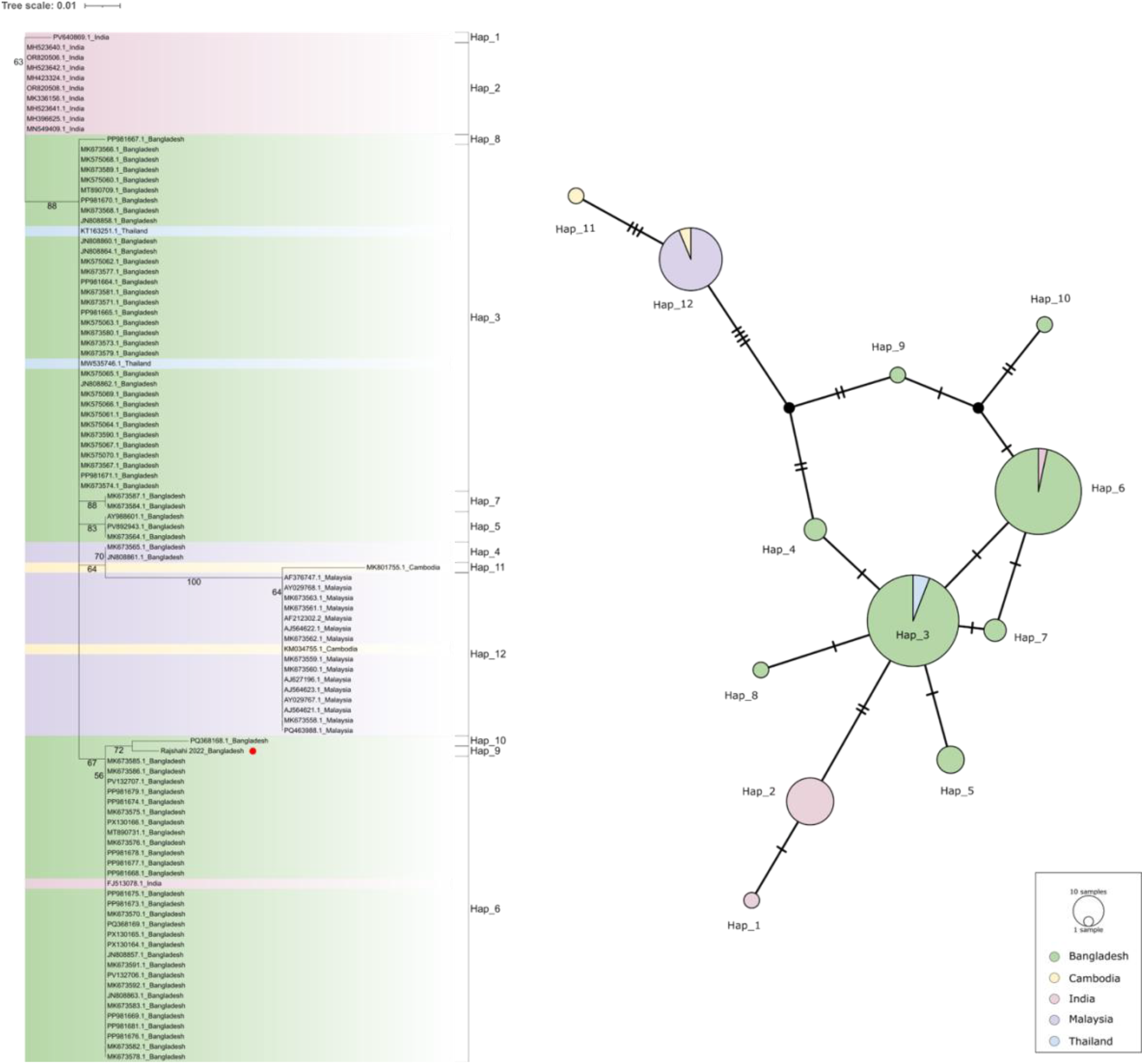
Haplotype-network analysis of the partial nucleocapsid sequence of NiV positive sample from 2022, Bangladesh, compared to cognate sequences from the region.

Based on the phylogenetic analysis the sequence from 2025 falls within NiV Bangladesh sublineage 2, as part of a genetically coherent cluster that has been repeatedly detected among Bangladeshi NiV sequences (Fig. 7).

**Fig. 7:**
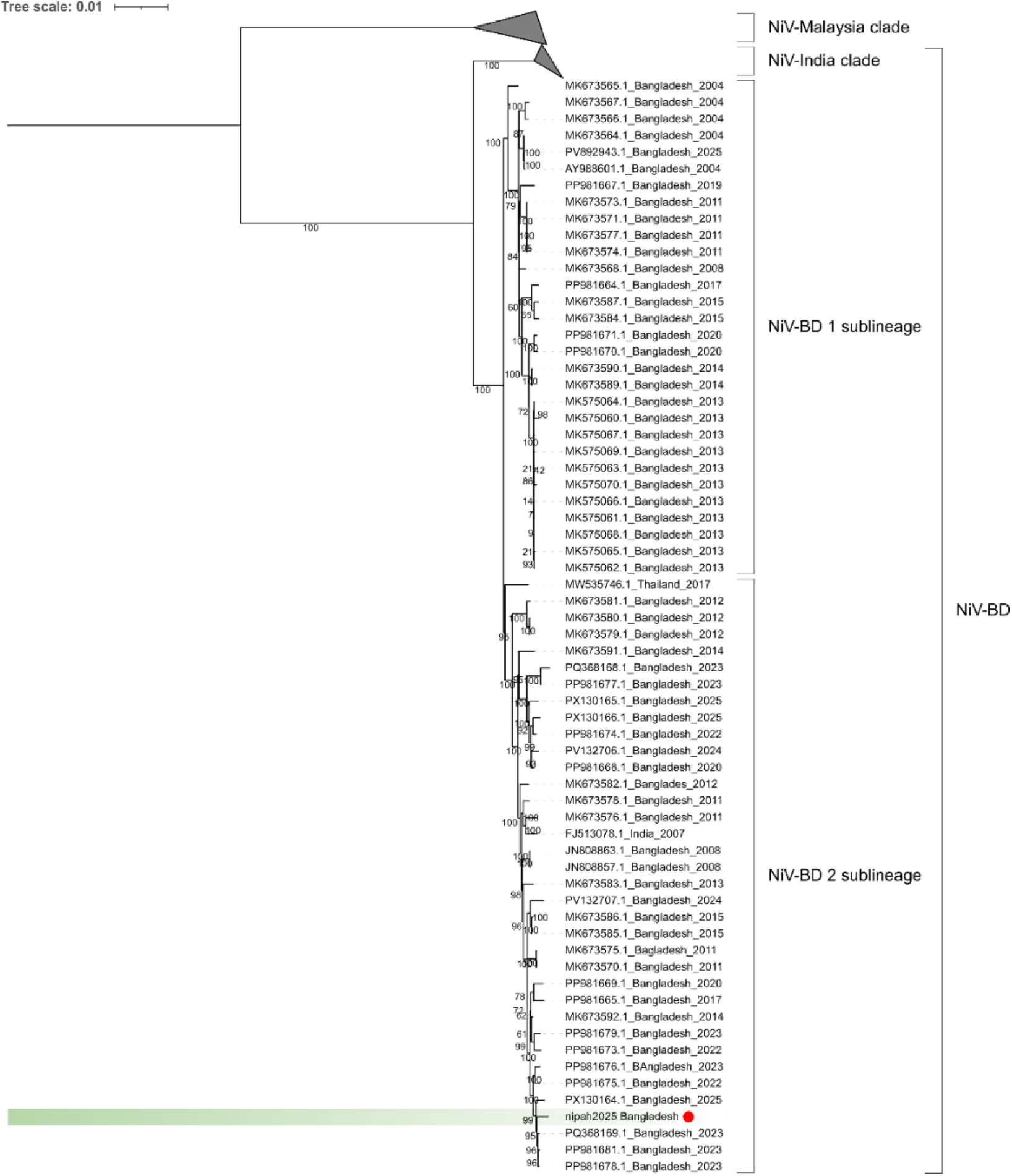
Maximum likelihood phylogenetic analysis of the partial nucleocapsid gene of NiV positive sample from 2025, Bangladesh.

## Discussion

Mobile surveillance platforms are increasingly recognized as essential tools for outbreak preparedness, response, and prevention. In recent years, several successful initiatives have demonstrated the feasibility of mobile laboratories for real-time or near real-time genomic surveillance of pathogens in both human and animal populations ^15^. As a relevant example, during the recent Nipah outbreak in West Bengal, the WHO reported that Indian authorities deployed a mobile Biosafety level-3 (BSL-3) laboratory to support outbreak response activities^10^. This mobile laboratory setup was previously published in detail. Although this is more of a technological demonstration than a technical one, it is the first published milestone specifically for the fight against Nipah, and it has been actively used ever since ^13^. In case of zoonotic diseases, where the natural reservoir and possible spillover mechanisms are known, these mobile laboratory solutions can serve as an integrated One Health surveillance tool. This approach opens the possibility for early notification of both veterinary and public health authorities. In developing countries or rural areas where access to molecular diagnostic infrastructure is limited by financial or infrastructural constraints, these solutions have a real potential for multiple purposes in outbreak prevention and response. The need for diagnostic capabilities, especially in one health scenarios, is increasingly in demand. While the development of therapeutic options and the exploration of related virus parameters receive significant attention, the diagnostic technology gap persists^20^. The use and development of mobile laboratory solutions is becoming increasingly common, yet there is a great lack of validated protocols in the literature that make sample inactivation and analytical processes safe and repeatable and paving the way for a more standardized way of basic operation. We followed this developmental pathway by establishing novel field-applicable methods, validating them under laboratory conditions, and subsequently demonstrating their integration and application during field surveillance activities.

Proper virus inactivation protocols are crucial elements in non-laboratory environments but are often missing in related publications. As the Nipah research portfolio becomes increasingly diversified, from epidemiological studies to ecological surveys to the development of new diagnostic procedures ^7,12,21–23^, the importance of protocols that enable appropriate inactivation of the virus under different laboratory and non-laboratory conditions is also increasing. This is crucial not only for mobile laboratory procedures but also in areas where adequate biosafety-classified laboratory space is unavailable to process primary Nipah virus samples. This is particularly important in developing countries or underdeveloped areas, so the results of our study in this regard are expected to increase responsiveness in future studies. While virus inactivation using TRI-Reagent was not specifically validated for the sample matrices used in this surveillance, the efficacy of this approach is strongly supported by existing literature ^24–27^. This also confirms the usefulness of Trizol-based inactivation solutions for similar studies.

Following the study’s concept of developing new methods and validating them in the laboratory, we also aimed to demonstrate their applicability in real-world settings. For this purpose, we decided to collect roost samples from bat colonies in NiV-endemic regions. Given the conservation status of bats and their declining populations, we believe this approach is essential, as it is non-invasive and causes minimal to no stress to the animals while still supporting epidemiological research. During validation, all field sampling was conducted using the developed mobile laboratory protocols, including inactivation of virus samples, PCR testing, and sequencing of positive samples. In addition to the many advantages of the method, it is also advantageous from the perspective of the Nagoya Protocol, as the full analysis of the samples is performed within 24 hours of collection at the sample collection site, and the resulting genomic data remains within the country.

The sequencing method presented in this publication is not the first of its kind. Recently, an amplicon-based sequencing approach validated on diverse human clinical samples has been published and adapted to the Bangladesh strain of Nipah virus ^22^. Furthermore, a recently published protocol describes a sequencing method for Nipah virus based on Nanopore technology ^28^. However, we developed a multi-layered, dual primer set design to increase genome coverage and improve performance across a wider range of target regions, including partially degraded or fragmented samples. A limitation of our study is that validation was performed primarily under laboratory conditions using the Nipah virus Malaysia strain, with additional testing on a single field-collected sample from Bangladesh. Nevertheless, the main objective of this work was to establish an integrated, field-tested mobile laboratory workflow that remains flexible and allows the incorporation of alternative sequencing approaches as needed. The Nipah Malaysia strain was used to validate the developed method, with successful genome amplification, suggesting that the sequencing method is capable of partial or complete amplification of diverse strains. However, this requires further laboratory and field validation. Other methods based on this concept are currently available, any of which can be integrated into the workflow we have published here.

The method’s significance is relevant to genomic epidemiological analysis of human epidemics, wildlife monitoring, and other situations where rapid genomic analysis may be required. In addition to general phylogenetic analysis, monitoring the variability and mutations of circulating Nipah strains may also be crucial to aid antiviral treatment, diagnostic procedures, or vaccine development.

As a result of phylogenetic and haplotype analyses, the positive samples from 2022 and 2025 can also be classified as the NiV Bangladesh 2 sublineage, which the original descriptive publication refers to as a sporadic sublineage. Overall, the placement of the novel sequence from 2025 supports the view that sublineage 2 remains actively maintained over time, with sequential sampling capturing a lineage that is genetically stable but continuously evolving – a typical genetic drift of an endemic lineage. The 2025 isolate is evolutionarily linked to previously circulating strains, reflecting ongoing persistence and gradual genetic turnover. This pattern is consistent with ongoing microevolution, potentially driven by local transmission dynamics, host adaptation, or stochastic accumulation of mutations rather than long-term independent evolution. These findings, although the analysis was made on a smaller genetic section, can still be interpreted stably for the 2022 sample. Thus, it can be stated that we succeeded in detecting the endemic NiV BD 2 sublineage circulating in the bat reservoir during the demonstration of the method and in generating genetic data in the field.

In Bangladesh, over the past decades, substantial research efforts have focused not only on human cases but also on monitoring flying foxes, which are now recognized as the natural reservoirs of Nipah virus. Surveillance studies in bats have revealed key ecological factors shaping Nipah virus dynamics within its reservoir host^12^. To improve the speed and accuracy of surveillance and better understand the emergence of outbreaks in relation to human infections, parallel bat monitoring activities were also conducted. In one such study, the authors identified a genetic link between Nipah virus strains circulating in local bat populations and those responsible for human cases, providing direct evidence of spillover from bats to humans ^21^.

Although these approaches have been highly valuable in advancing our understanding of the virus, from an epidemiological perspective, most have relied on classical surveillance strategies or reactive outbreak investigations. Despite the widely recognized need - highlighted by WHO and other expert bodies - there is still no established early warning system capable of supporting realistic risk assessment prior to outbreak emergence. Here, we present a fully developed and field-tested mobile laboratory workflow, representing the first published procedure that enables rapid deployment and integrated, complex surveillance under real-world conditions.

In conclusion, the development of rapid on-site surveillance techniques, starting with a validated inactivation protocol, followed by on-site real-time RT-PCR screening and amplicon-based sequencing, represents a significant advancement in the early detection and monitoring of NiV. This approach facilitates swift responses in the early stages of human cases and contact tracing, as well as the identification of virus presence in bat colonies before human cases emerge, bolstering outbreak prevention efforts. Moreover, the portability and ease of use of these mobile sequencing technologies are promising to vaccine development and enhance our understanding of NiV evolution in both natural and outbreak settings. Overall, these advancements highlight the crucial role of proactive surveillance measures in mitigating the risk of NiV outbreaks and highlight the importance of continued development in innovative technologies for rapid pathogen detection and characterization in field settings.

## Data Availability

All data produced in the present work are contained in the manuscript

## Contributors

Conceptualization: MNI, SAK, ZL, NCD, GK

Data curation: ZL, ÁÁ, BZ

Formal analysis: ÁÁ, BZ, JAUJ

Funding acquisition: GK

Investigation: MNI, SAK, ZL, SA, MSI, ASMRJ, JAUJ, GK

Methodology: ZL, ÁÁ, BZ, SZ, KL, GK

Project administration: MNI, SA, AAMB, MJUS, MHI, GK, SC

Supervision: SAK, MJUS, MHI, NCD, GK, SC

Validation: ZL, ÁÁ, KL

Visualization: ZL, ÁÁ, BZ, JAUJ

Data interpretation: ZL, ÁÁ, GK

Writing - original draft: ZL, GK

Writing - review & editing: MNI, SAK, ZL, ÁÁ, SA, AAMB, BZ, SZ, JAUJ, KB, SC

## Acknowledgements

The authors thank the Bangladesh Forest Department for granting research permissions and providing logistical support. We gratefully acknowledge the Department of Livestock Services, along with the respective District and Upazila Livestock Officers, for their local assistance and for providing the facilities necessary to establish the mobile laboratory. Finally, we extend our thanks to Gábor E. Tóth, Soumya Khan, and Monsur Sarkar and son for their technical support and fieldwork assistance. Two colleagues were supported by the 2025-2.1.1-EKÖP funding scheme by the Ministry of Culture and Innovation, National Fund for Research, Development and Innovation, under the University Research Grant Programme Á.Á.: EKÖP-25-3-I-PTE-669, and Z.L.: EKÖP-25-4-II-PTE-470.

## References

1 Aditi, Shariff M. Nipah virus infection: A review. Epidemiol Infect 2019; 147: e95.

2 Chua KB, Bellini WJ, Rota PA, et al. Nipah Virus: A Recently Emergent Deadly Paramyxovirus. Science (80-) 2000; 288: 1432–5.

3 Hsu VP, Hossain MJ, Parashar UD, et al. Nipah Virus Encephalitis Reemergence, Bangladesh. Emerg Infect Dis 2004; 10: 2082–7.

4 Sahay RR, Yadav PD, Gupta N, et al. Experiential learnings from the Nipah virus outbreaks in Kerala towards containment of infectious public health emergencies in India. Epidemiol Infect 2020; 148: e90.

5 Jacob John T, Gupta N, Vasant Murhekar M. Nipah virus infection in humans in Kerala, India: Hypothesis of air-borne transmission. Indian J Med Res 2025; 161: 567.

6 Yadav PD, Baid K, Patil DY, et al. A One Health approach to understanding and managing Nipah virus outbreaks. Nat Microbiol 2025; 10: 1272–81.

7 Nikolay B, Salje H, Hossain MJ, et al. Transmission of Nipah Virus — 14 Years of Investigations in Bangladesh. N Engl J Med 2019; 380: 1804–14.

8 Nazmunnahar, Ahmed I, Roknuzzaman ASM, Islam MR. The recent Nipah virus outbreak in Bangladesh could be a threat for global public health: A brief report. Heal Sci Reports 2023; 6. DOI:10.1002/hsr2.1423.

9 Khan S, Akbar SMF, Mahtab M Al, et al. Twenty-five years of Nipah outbreaks in Southeast Asia: A persistent threat to global health. IJID Reg 2024; 13: 100434.

10 WHO. Disease Outbreak News: Nipah virus infection in India. World Heal. Organ. 2026. https://www.who.int/emergencies/disease-outbreak-news/item/2026-DON593 (accessed Jan 30, 2026).

11 WHO. WHO South-East Asia Regional Strategy for the prevention and control of Nipah virus infection 2023–2030. Printed in India, 2023.

12 Epstein JH, Anthony SJ, Islam A, et al. Nipah virus dynamics in bats and implications for spillover to humans. Proc Natl Acad Sci 2020; 117: 29190–201.

13 Sahay RR, Patil DY, Shete AM, et al. Rapidly deployable mobile BSL-3 laboratory: a response to the Nipah virus outbreak in Kozhikode, Kerala, India, 2023. Pathog Glob Health 2025; : 1–10.

14 Mackay IM, Northill JA. Nipah virus real-time RT-PCR (NiV-TM2018). 2019. DOI:10.17504/protocols.io.rs5d6g6.

15 Quick J, Loman NJ, Duraffour S, et al. Real-time, portable genome sequencing for Ebola surveillance. Nature 2016; 530: 228–32.

16 Li H. Minimap2: pairwise alignment for nucleotide sequences. Bioinformatics 2018; 34: 3094–100.

17 Katoh K, Standley DM. MAFFT Multiple Sequence Alignment Software Version 7: Improvements in Performance and Usability. Mol Biol Evol 2013; 30: 772–80.

18 Minh BQ, Schmidt HA, Chernomor O, et al. IQ-TREE 2: New Models and Efficient Methods for Phylogenetic Inference in the Genomic Era. Mol Biol Evol 2020; 37: 1530–4.

19 Letunic I, Bork P. Interactive Tree of Life (iTOL) v6: recent updates to the phylogenetic tree display and annotation tool. Nucleic Acids Res 2024; 52: W78–82.

20 Asokan S, Luke MS, Atiyah HM, et al. Nipah virus as a pandemic threat: Current knowledge, diagnostic gaps, and future research priorities. Diagn Microbiol Infect Dis 2026; 114: 117141.

21 McKee CD, Islam A, Rahman MZ, et al. Nipah Virus Detection at Bat Roosts after Spillover Events, Bangladesh, 2012–2019. Emerg Infect Dis 2022; 28: 1384–92.

22 Rahman MM, Miah M, Hossain ME, et al. Development of a culture-independent whole-genome sequencing of Nipah virus using the MinION Oxford Nanopore platform. Microbiol Spectr 2025; 13. DOI:10.1128/spectrum.02492-24.

23 Rahman MM, Rahman S, Rahim S, et al. Mapping the geodispersal and evolutionary dynamics of regional Nipah virus strain in Bangladesh. Microbiol Spectr 2026; published online March 12. DOI:10.1128/spectrum.02319-25.

24 Olejnik J, Hume AJ, Ross SJ, et al. Art of the Kill: Designing and Testing Viral Inactivation Procedures for Highly Pathogenic Negative Sense RNA Viruses. Pathogens 2023; 12: 952.

25 Retterer C, Kenny T, Zamani R, et al. Strategies for Validation of Inactivation of Viruses with Trizol® LS and Formalin Solutions. Appl Biosaf 2020; 25: 74–82.

26 Silva-Ayala D, Griffiths A. Validation of Chemical Inactivation Protocols for Henipavirus-Infected Tissue Samples. Viruses 2026; 18: 81.

27 Widerspick L, Vázquez CA, Niemetz L, et al. Inactivation Methods for Experimental Nipah Virus Infection. Viruses 2022; 14: 1052.

28 Ou TP, Pum L, Rath S, et al. Sequencing of Nipah Virus (NiV) Using Oxford Nanopore MinION v1. 2026; published online Feb 2. DOI:10.17504/protocols.io.6qpvryr5pgmk/v1.

