## Supplements for "Development of an early warning system for Nipah outbreak prevention: on-site inactivation, PCR surveillance and sequencing in Bangladesh"

**Appendix A Supplementary data (1)**

**Supplementary tables (S1-S7) and figures (S1)**

**Supplementary Table S1** Primer sets were designed with PrimalScheme for the following genomes obtained from GenBank database.

MK575063.1

MK673580.1

MK673581.1

MK673579.1

MK673591.1

MK673586.1

FJ513078.1

MK673576.1

MK673582.1

MK673583.1

MK673578.1

JN808863.1

JN808857.1

MK673588.1

MK673575.1

MK673570.1

MK673592.1

MK673585.1

MW535746.1

MK673587.1

MK673584.1

MK673565.1

MK575060.1

MK575070.1

MK575065.1

MK575062.1

MK575067.1

MK575069.1

MK575061.1

MK575064.1

MK575068.1

MK575066.1

MK673590.1

MK673589.1

JN808864.1

MK673577.1

MK673571.1

MK673573.1

MK673574.1

MK673572.1

MK673567.1

MK673566.1

MK673568.1

MK673564.1

AY988601.1

**Supplementary Table S2** Primer sequences and their positions in reference to genome AY988601.1

| **primer** | **sequence** | **position** |
| --- | --- | --- |
| nipah_600_1_LEFT | GCTAGTTTTAGGAGTTATCAATCTAAGTTAGG | 137-168 |
| nipah_600_1_RIGHT | AGAGCAAAGAACGGATTGACTCT | 716-738 |
| nipah_600_2_LEFT | TGCCAAAGCAGTTACAGCTCC | 640-660 |
| nipah_600_2_RIGHT | TCTGATCAATCCCTCCAGCATG | 1220-1241 |
| nipah_600_3_LEFT | TGACAGGTCTATGGGGGCATTA | 1135-1156 |
| nipah_600_3_RIGHT | TTGTAGAATATTGAAAATAAGCGTCACAC | 1706-1734 |
| nipah_600_4_LEFT | GCAATCAGCAACAAAGATCAAAGG | 1613-1636 |
| nipah_600_4_RIGHT | TCATTTATGTGAGCATTACCTAGTAGATCT | 2195-2224 |
| nipah_600_5_LEFT | AGCTATTGTCTTTGCACTGGAGT | 2100-2122 |
| nipah_600_5_RIGHT | GTCATCTGAACCTTCTGCCCAG | 2696-2717 |
| nipah_600_6_LEFT | TGGAGGTGTTGAAAGAAGAAGCT | 2606-2628 |
| nipah_600_6_RIGHT | CCTCTTCAGGAGAAATGCCGAC | 3192-3213 |
| nipah_600_7_LEFT | ACACATCAGATGACGAAGAGGC | 3118-3139 |
| nipah_600_7_RIGHT | ATCCTCGAACATGCCGGGTT | 3686-3705 |
| nipah_600_8_LEFT | AGGAACTCCGATGCCAAAATCC | 3581-3602 |
| nipah_600_8_RIGHT | AGCTCAGGATTACTTTTGCCTTTTC | 4162-4186 |
| nipah_600_9_LEFT | CAGGGTTCTGGCCAAAACCAA | 4067-4087 |
| nipah_600_9_RIGHT | TCAAGTACTGTGTTATCGCCCG | 4661-4682 |
| nipah_600_10_LEFT | ACATGTAGACAGTAGTTTCAGGTCA | 4582-4606 |
| nipah_600_10_RIGHT | CCATTCTCCCAAGAACTAGGGC | 5166-5187 |
| nipah_600_11_LEFT | AAATTCACCAGCCCTTGCCAAG | 5073-5094 |
| nipah_600_11_RIGHT | ACTCAAGCATAGTTCGTGGAATCA | 5670-5693 |
| nipah_600_12_LEFT | TGCCGAAACGTTGATCAGATACA | 5576-5598 |
| nipah_600_12_RIGHT | TTTGAAAAGGATTCTACTTAGCCCTTT | 6157-6183 |
| nipah_600_13_LEFT | AACCTGGAACAACAGCTGTGAG | 6043-6064 |
| nipah_600_13_RIGHT | CTGCCATTGTCGAGTCTTACGG | 6645-6666 |
| nipah_600_14_LEFT | AGGTCGCGGGAATACAAACAAA | 6533-6554 |
| nipah_600_14_RIGHT | AGCTTCATTAGTTGATTCTATGCTGC | 7105-7130 |
| nipah_600_15_LEFT | GCTCAAATTACTGCAGGTGTAGC | 7035-7057 |
| nipah_600_15_RIGHT | TGTGTTCCTTACCAATATGAAATTTGGG | 7595-7622 |
| nipah_600_16_LEFT | TGACTGAAATCCAACAGGCCTA | 7513-7534 |
| nipah_600_16_RIGHT | CGGGTTAACAGTATCAAGGAGTCG | 8094-8117 |
| nipah_600_17_LEFT | TGTCAAACAACAGGTAGGGCAA | 7839-7860 |
| nipah_600_17_RIGHT | AAAGTTGCAACATTCCTATCTCAAGT | 8393-8418 |
| nipah_600_18_LEFT | TGAATTACATCAAAGAGATCAAAGGACT | 8316-8343 |
| nipah_600_18_RIGHT | TGTGCCAGCAATGCTTTCTTCT | 8879-8900 |
| nipah_600_19_LEFT | TCACATTATAGGAGTGAGCCACAA | 8836-8859 |
| nipah_600_19_RIGHT | AAGGGAGTGGGTTAGGACAAGA | 9429-9450 |
| nipah_600_20_LEFT | AGATCAGCCAGTCAACTGCAAG | 9337-9358 |
| nipah_600_20_RIGHT | CGTAAGGCAAATTGATGTTGATTGT | 9937-9961 |
| nipah_600_21_LEFT | AATAGCACCTACTGGTCCGGAT | 9864-9885 |
| nipah_600_21_RIGHT | TGGGCACTTGTTGAATCTAGGG | 10427-10448 |
| nipah_600_22_LEFT | GGAGATGTCCAAACAGTTAACCCT | 10347-10370 |
| nipah_600_22_RIGHT | TCAAGATTATTTATTGACTCGGTCTCC | 10926-10952 |
| nipah_600_23_LEFT | TGTCATAATTACATAAGTCTTCAGCTGAA | 10835-10863 |
| nipah_600_23_RIGHT | GTCAGATATGGATAATTCATCGGCC | 11420-11444 |
| nipah_600_24_LEFT | ACAGACTGAAGACGTTATTACCTATTTT | 11318-11345 |
| nipah_600_24_RIGHT | ACCATTTAGATTGACTCATATTCTTCCCT | 11888-11916 |
| nipah_600_25_LEFT | TGCTACAAAACATCACTAGAAATCTCA | 11800-11826 |
| nipah_600_25_RIGHT | AGCCACACTCACTCAATTCCTG | 12378-12399 |
| nipah_600_26_LEFT | CCCTTAGTTCTTGCACTACTCCA | 12291-12313 |
| nipah_600_26_RIGHT | TGGGGTATAGCTCAGCACTTCT | 12884-12905 |
| nipah_600_27_LEFT | TCATGGAGCTGAAATTAGACAGTGA | 12790-12814 |
| nipah_600_27_RIGHT | ATGATAGACCCCGTCAGCTTGG | 13373-13394 |
| nipah_600_28_LEFT | TCCAATGCTTTAAAGGAGTTACCAC | 13285-13309 |
| nipah_600_28_RIGHT | TCCTCGTGTTTGTCTCATAGGC | 13869-13890 |
| nipah_600_29_LEFT | TCCTAAAGGCGGTATTGAAGGAT | 13796-13818 |
| nipah_600_29_RIGHT | TAGGTGCTGGGATTAATGCTGC | 14382-14403 |
| nipah_600_30_LEFT | GCAACTCCTTATATCAACTGAGTTTAGT | 14288-14315 |
| nipah_600_30_RIGHT | TCTGGAAACTAACCTTGGCTGAA | 14878-14900 |
| nipah_600_31_LEFT | GGACAATTCATTGACAGGTGCC | 14786-14807 |
| nipah_600_31_RIGHT | TGAGACGCTAGGTACCAAGCTT | 15367-15388 |
| nipah_600_32_LEFT | GAGACCGACTAAAGCCTTACGT | 15281-15302 |
| nipah_600_32_RIGHT | CCTCAGTTGACCATAAAGGGAAGT | 15862-15885 |
| nipah_600_33_LEFT | TCTTATATATGATCCAGATCCTGTTTCAGA | 15782-15811 |
| nipah_600_33_RIGHT | AGTTTCATCCTGTTCGGCGATT | 16361-16382 |
| nipah_600_34_LEFT | GCTGTCTCAAAATCTCCTTGTAACA | 16283-16307 |
| nipah_600_34_RIGHT | GATGATGAGTTGAGGCCTATCCTT | 16847-16870 |
| nipah_600_35_LEFT | AGGAAATCCTGTCTATGAACACGA | 16759-16782 |
| nipah_600_35_RIGHT | AATCCGGGGGTGTATGCTATCT | 17344-17365 |
| nipah_600_36_LEFT | AGAAGAGATACTAGTAGAACATTCTCATCT | 17264-17293 |
| nipah_600_36_RIGHT | TGGATACGGTTCCAAATGATGTGA | 17832-17855 |
| nipah_600_37_LEFT | ACAGTTCCATGAAGATCTAAAGAAATACT | 17597-17625 |
| nipah_600_37_RIGHT | GCACAAATTGTCGGTCGGTTC | 18171-18191 |
| nipah_900_1_LEFT | CGGCTAGTTTTAGGAGTTATCAATCTAA | 135-162 |
| nipah_900_1_RIGHT | GCTCTTGGGCCAATTTCTCTGT | 1014-1035 |
| nipah_900_2_LEFT | TCGCAACCATCAGATTTGGGTT | 915-936 |
| nipah_900_2_RIGHT | TTAATGTAATTGGTCCCTTAGTGTTGT | 1770-1796 |
| nipah_900_3_LEFT | ACCAAGATCTCAAACCCACTCAA | 1656-1678 |
| nipah_900_3_RIGHT | TCCCATGCTTTTGTTCGGTCTT | 2521-2542 |
| nipah_900_4_LEFT | AAATTGGAACTAGTTAATGATGGCCT | 2412-2437 |
| nipah_900_4_RIGHT | TTTGGAGGCTGTCTCGAGTTGA | 3276-3297 |
| nipah_900_5_LEFT | TGCAGGGAGTTCAAGTGAAGTG | 3167-3188 |
| nipah_900_5_RIGHT | TTTTGGCCAGAACCCTGTCAAT | 4062-4083 |
| nipah_900_6_LEFT | TGCGAAGAATCAGTTTTGATGGG | 3939-3961 |
| nipah_900_6_RIGHT | TCTGCTCCATGCTGATATCAGATT | 4819-4842 |
| nipah_900_7_LEFT | TGAAGTGTATGTAGCATGATCAAATTACT | 4694-4722 |
| nipah_900_7_RIGHT | TTCTCAGGGCTTGATGCTTGTC | 5603-5624 |
| nipah_900_8_LEFT | AAATCACCTTGTCCCGTGGAAG | 5512-5533 |
| nipah_900_8_RIGHT | GAGCTTGGCTCCTAAGTTTTTCAT | 6359-6382 |
| nipah_900_9_LEFT | ACATCACGCAGGACTATTTACAAAA | 6225-6249 |
| nipah_900_9_RIGHT | TCTTTTCTGCAGTCTCTTGAAGCT | 7138-7161 |
| nipah_900_10_LEFT | GCTCAAATTACTGCAGGTGTAGC | 7035-7057 |
| nipah_900_10_RIGHT | CCAAGCTGATGATCACATTACCG | 7925-7947 |
| nipah_900_11_LEFT | TGACTGAAATCCAACAGGCCTA | 7513-7534 |
| nipah_900_11_RIGHT | TGCAACATTCCTATCTCAAGTATCCA | 8388-8413 |
| nipah_900_12_LEFT | TGTACTATCAATTGCATCATTGTGTATAGG | 8153-8182 |
| nipah_900_12_RIGHT | TCATTTATTTTCTTAATGTCCATGGTTCC | 9036-9064 |
| nipah_900_13_LEFT | TGCCGACAGAAAGCAAGAAAGT | 8950-8971 |
| nipah_900_13_RIGHT | TCCAACAACTGACACTGCACAA | 9830-9851 |
| nipah_900_14_LEFT | ATGACTAACGTCTGGACCCCAT | 9747-9768 |
| nipah_900_14_RIGHT | TTTTGTGCATTGGTGTCCTCGG | 10606-10627 |
| nipah_900_15_LEFT | CGGGTGTATTCCTTGACAGCA | 10513-10533 |
| nipah_900_15_RIGHT | TTGGTAGCCAATTTTTAGGAGAATTAGA | 11391-11408 |
| nipah_900_16_LEFT | CCCAGGTCCTTGATTGTGCTAA | 11269-11290 |
| nipah_900_16_RIGHT | TGTCTCCATCATCATCCTCCCT | 12146-12167 |
| nipah_900_17_LEFT | TCCTGAATCCAAATCTTATCTGCATTT | 12043-12069 |
| nipah_900_17_RIGHT | TCAACCAATCTTCTTGGCTCGG | 12916-12937 |
| nipah_900_18_LEFT | ATGAAAGATAAAGCCTTGTCTCCTATC | 12828-12854 |
| nipah_900_18_RIGHT | GGGCAATTAGGATCCGCAACAT | 13711-13732 |
| nipah_900_19_LEFT | TGGCCATATTTGCTGAACGTCT | 13621-13642 |
| nipah_900_19_RIGHT | CGCTTTCAGTCATAATGCTGTGG | 14501-14523 |
| nipah_900_20_LEFT | GCAGCATTAATCCCAGCACCTA | 14382-14403 |
| nipah_900_20_RIGHT | ACGTAAGGCTTTAGTCGGTCTC | 15281-15302 |
| nipah_900_21_LEFT | CCTAGGGATTCCCAATTAGATCAGG | 15171-15195 |
| nipah_900_21_RIGHT | TTGTCCAAGATAAAGTGCAAACAGT | 16037-16061 |
| nipah_900_22_LEFT | GGCTGACAAGGATGTTTTAAAGCA | 15944-15967 |
| nipah_900_22_RIGHT | CCTTCTATATTTGTGATTCTCCACCG | 16825-16850 |
| nipah_900_23_LEFT | CCTCCTATCAAAACAGGGGTGT | 16704-16725 |
| nipah_900_23_RIGHT | GATCTTCATGGAACTGTTTAGATTGAGA | 17586-17613 |
| nipah_900_24_LEFT | ATCGCGATAAATGTTATGATGGAGG | 17301-17325 |
| nipah_900_24_RIGHT | AAATTGTCGGTCGGTTCTGGA | 18167-18187 |
| nipah_0_LEFT | ATATATAACGTATTTCTAAAACTTAGGAACCA | 32-63 |
| nipah_0_LEFT_ALT | CAAGGGAAAATATGGATACGTTAAA | 7-31 |
| nipah_END_RIGHT | ACCGAACAAGGGTAAAGAAGAATCGTT | 18226-18252 |
| nipah_END_RIGHT_ALT | GGGTAAAGAAGAATCGTTATTAAGTT | 18218-18243 |

**Supplementary Table S3** Coverage and genome completeness of the sequenced samples

| **Sample** | **Average coverage per base** | **Complete bases** | **Completeness: percentage** | **reference length** |
| --- | --- | --- | --- | --- |
| malaysia_Ct 25,27 | 24760 | 17709 | 97 | 18246 |
| malaysia_Ct 31,27 | 24113 | 16242 | 89 | 18246 |
| malaysia_Ct 34,2 | 1697 | 11147 | 61 | 18246 |
| malaysia_Ct 34,99 | 239 | 9936 | 54 | 18246 |
| sample from 2025, Bangladesh | 3971 | 5537 | 30 | 18126 |

**Standard curve and copy number calculation of samples**

The amplified positive control from the qRT-PCR reaction was run on agarose gel, to make sure nothing aspecific was present in the PCR product. Then the band was excised and purified with NucleoSpin Gel and PCR Clean-up kit (Macherey-Nagel, Germany). The concentration of the elute was measured with Qubit™ 1X dsDNA HS Assay Kit (Invitrogen, USA) and was measured as 4,44 ng/µL. From this the copy number of this product was calculated as 33,6 * 10^9 copies/µL. From this concentrated product a 10-fold serial dilution of samples was established, from 1*10^8 to 1*10^1 copies/ µl. The dilutes were then run with the same qRT-PCR described in the Materials and methods section. Based on the resulting standard curve, we calculated the copy number of samples.

**Supplementary Fig. S1** Standard curve (Ct value in relation to logarithmized copies/µl value): y = -3,7324x + 37,303; where y is the Ct value.

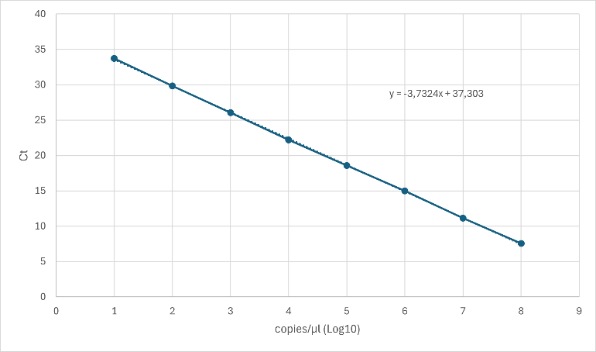

**Supplementary Table S4** Serial dilution of PCR product for standard curve

| **copy_number_log10** | **copy_number** | **Ct** |
| --- | --- | --- |
| 8 | 100000000 | 7,54 |
| 7 | 10000000 | 11,13 |
| 6 | 1000000 | 15 |
| 5 | 100000 | 18,59 |
| 4 | 10000 | 22,19 |
| 3 | 1000 | 26,06 |
| 2 | 100 | 29,84 |
| 1 | 10 | 33,71 |

**Supplementary Table S5** Serial dilution of Malaysia strain Nipah isolate RNA

| **copy_number_log10** | **copy_number** | **Ct** |
| --- | --- | --- |
| 3,22393 | 1675 | 25,27 |
| 1,61639 | 41 | 31,27 |
| 0,83137 | 7 | 34,2 |
| 0,61971 | 4 | 34,99 |

**Supplementary Table S6** Copy number of the 2025 Bangladesh Nipah sample

| **copy_number_log10** | **copy_number** | **Ct** |
| --- | --- | --- |
| 0,61703 | 4 | 35 |

**Supplementary Table S7** Metadata for haplotype analysis

| **Haplotype** | **Frequency** | **# of Countries** | **Name of countries** | **Accession numbers** |
| --- | --- | --- | --- | --- |
| Hap_1 | 1 | 1 | India | PV640869.1 |
| Hap_2 | 9 | 1 | India | MN549409.1 MH523640.1 MH396625.1 MH523642.1 MH423324.1 MH523641.1 MK336156.1 OR820506.1 OR820508.1 |
| Hap_3 | 34 | 2 | Bangladesh, Thailand | MK673574.1 MK673566.1 MK673568.1 MK673567.1 MK673573.1 MK673571.1 MK575070.1 MK575060.1 MK575061.1 MK575062.1 MK575063.1 MK575064.1 MK575065.1 MK575066.1 MK575067.1 MK575068.1 MK575069.1 MK673580.1 MK673581.1 MW535746.1 MK673577.1 MK673590.1 MK673589.1 MK673579.1 PP981670.1 PP981671.1 JN808858.1 JN808860.1 JN808862.1 JN808864.1 KT163251.1 MT890709.1 PP981664.1 PP981665.1 |
| Hap_4 | 2 | 1 | Bangladesh | JN808861.1 MK673565.1 |
| Hap_5 | 3 | 1 | Bangladesh | MK673564.1 AY988601.1 PV892943.1 |
| Hap_6 | 30 | 2 | Bangladesh, India | MK673578.1 MK673585.1 MK673582.1 MK673576.1 MK673575.1 MK673586.1 MK673570.1 JN808857.1 JN808863.1 PP981677.1 PP981668.1 PP981674.1 PP981679.1 PP981675.1 PP981673.1 PP981676.1 PP981669.1 PQ368169.1 PP981678.1 PP981681.1 PX130164.1 FJ513078.1 MK673583.1 MK673592.1 MT890731.1 PV132707.1 PX130166.1 PX130165.1 PV132706.1 MK673591.1 |
| Hap_7 | 2 | 1 | Bangladesh | MK673584.1 MK673587.1 |
| Hap_8 | 1 | 1 | Bangladesh | PP981667.1 |
| Hap_9 | 1 | 1 | Bangladesh | Rajshahi_2022 |
| Hap_10 | 1 | 1 | Bangladesh | PQ368168.1 |
| Hap_11 | 1 | 1 | Cambodia | MK801755.1 |
| Hap_12 | 16 | 2 | Cambodia, Malaysia | PQ463988.1 AF376747.1 AJ627196.1 AY029768.1 KM034755.1 MK673562.1 MK673558.1 MK673559.1 MK673560.1 MK673561.1 MK673563.1 AF212302.2 AJ564621.1 AJ564622.1 AJ564623.1 AY029767.1 |
